# Previewing gaze behaviours predict gait speed, risky stepping strategies and cognitive dual-task cost in older adults

**DOI:** 10.64898/2026.11.16.26368206

**Authors:** Olivia McVeigh-Mellor, Adam J. Cocks, Liam O’Reilly, Dominic Orth, Adam Bruton, Toby Ellmers, Elmar Kal

## Abstract

**Background:** Previous work demonstrated that route previewing improves adaptive gait performance in older adults. However, it is unknown if specific gaze behaviours during previewing are associated with these benefits. Specifically, little is known about how older adults visually scan future walking routes during a given previewing period and whether different gaze behaviours are associated with subsequent walking performance.

**Methods:** Gaze and gait data were collected from 29 older adults (*M*_age_ = 73.9, *SD* = 8.1 years; *n* = 19 female) during dual-task adaptive walking, preceded by a 10-second period in which they were instructed to preview their walking route. Eye-tracking measures across defined areas of interest during previewing and subsequent gait measures were analysed using linear and generalized mixed-effects models to determine whether previewing gaze behaviours predicted gait outcomes and cognitive dual-task cost.

**Results:** Greater gaze sequence complexity (LZC_task) during previewing was associated with lower cognitive dual-task cost during walking, Larger gaze transition sizes predicted slower walking speed while greater duration of distal fixation allocation was associated with an increased likelihood of risky stepping behaviours. No previewing gaze behaviours predicted more than one gait outcome, and no significant relationships were observed between gaze behaviours and stepping errors.

**Conclusions:** Different aspects of visual previewing were associated with distinct locomotor and cognitive outcomes. Greater gaze-sequence complexity was associated with lower cognitive dual-task cost. In contrast, larger gaze transitions predicted slower walking. Together, these findings suggest that effective previewing may depend on integrating proximal and distal route information into a coherent prospective stepping plan, rather than simply looking further ahead. Understanding how older adults visually plan upcoming routes may help inform approaches to reducing dual-task-related impairments in adaptive walking.

## 1. Introduction

Adaptive walking relies on using visual information for both feedforward route planning and feedback-based control (Patla, 1997; Reynolds & Day, 2005; Matthis et al., 2016). Older adults appear increasingly reliant on visual input for stability and may perform walking tasks poorly with additional cognitive load that competes for cognitive resources (Pieruccini-Faria et al., 2014; Dominguez-Zamora et al., 2020). When dual-tasking, older adults show more stepping errors (Ellmers et al., 2016; Cocks et al., 2025) likely due to a reduction in attentional resources available for task-relevant visuo-motor control processes (Ellmers et al., 2016). Cognitive dual-tasking has previously been shown to shift gaze towards the immediate walking path and away from future stepping targets (Ellmers, Cocks, & Young, 2020) or other more distant environmental features potentially reducing visuo-motor planning (Zukowski et al., 2021). These findings highlight the importance of identifying strategies that help older adults maintain optimal gaze behaviour even under cognitive load, as this may support safer and more stable adaptive gait.

One such strategy is route previewing. Recent work from our group demonstrated that allowing older adults to preview an adaptive walking route before movement initiation mitigated the negative effects of dual-tasking on stepping performance (McVeigh-Mellor et al., 2026). These findings suggest that previewing reduced the cognitive load associated with online visual search, freeing up attentional resources that can be allocated to secondary task performance (and thereby attenuating dual-task interference). However, this previous study focused primarily on the behavioural consequences of previewing and did not examine participants’ gaze behaviour during the previewing period itself. Consequently, it remains unclear whether there are specific gaze behaviours through which previewing supports subsequent gait performance.

Previewing is thought to help older adults map environmental constraints and plan future actions in advance (Curzon-Jones and Hollands, 2018), but this has not yet been explicitly tested. If so, the benefits of previewing may depend not only on *where* older adults allocate gaze, but on *how* they scan future stepping constraints (e.g., how much they move their gaze between different areas of the walking path). Most locomotor eye-tracking studies describe visual behaviour by measuring where participants look and for how long for but fail to capture how gaze shifts between these. This sequence of fixations may provide additional insight into how visual information is gathered and organised, which may be particularly important before gait initiation (Amor et al., 2016). For example, gaze-sequence complexity may reflect how visual information is organised during exploration, with higher entropy indicating less predictable and potentially less structured visual sampling (Shiferaw et al., 2019). However, it is unknown whether the complexity of gaze fixation sequences during previewing, or other characteristics reflecting how visual information is sampled and distributed across the route, are associated with subsequent adaptive walking performance.

The present secondary analysis therefore examined whether specific gaze characteristics during route previewing predict subsequent adaptive gait and cognitive outcomes in older adults. We assessed three complementary aspects of visual exploration: (i) the overall organisation of scanning behaviour, indexed by gaze complexity and maximal gaze transition size; (ii) the extent of visual sampling across future route locations; and (iii) the allocation of visual attention towards distal route information, indexed by the proportion of fixation time on the latter half of the walkway

### Hypothesis

Smaller maximal gaze transitions and greater relative proportion of distal fixation duration towards the latter half of the walkway will improve subsequent walking performance.

## 2. Method

This analysis used gaze and gait data previously collected as part of a larger cross-sectional, experimental study (McVeigh-Mellor et al., 2026) that examines the effects of route previewing on adaptive walking and gaze behaviour under cognitive dual-task conditions in older adults. The study protocol was approved by Brunel University of London’s College of Health, Medicine and Life Sciences’ Research Ethics Committee (reference: 45431-A-Sep/2024-52891-1).

### 2.1 Participants

The study sample included twenty-nine community dwelling older adults aged 60–87 years (M_age_ = 73.9, SD = 8.1) who are a sub-sample from a larger cross-sectional study conducted by the research team (McVeigh-Mellor et al., 2026). Participants were recruited through the Brunel Older people’s Reference Group, a longstanding local public involvement and engagement group. Participants were eligible if they were aged 60 years or older, had neither a musculoskeletal condition affecting balance nor progressive neurological diagnosis, were able to walk unaided for 10 meters, stand unsupported for at least 2 minutes, and had (corrected-to) normal vision. Participants were excluded if the MoCA score was <22 (Carson et al., 2018).

### 2.2 Experimental Setup

Please see McVeigh-Mellor et al., (2026) for a comprehensive overview of the study protocol. The protocol was adapted from Cocks et al., (2025). Adaptive walking trials were conducted on an 8-metre instrumented walkway (Zeno Walkway system, ProtoKinetics, Havertown, PA, USA) marked with four coloured 10×10 cm stepping targets (red, blue, yellow and green) that defined different winding routes (see Figure 1). Each route was purposefully designed to have similar complexity. For each condition, all 4 coloured routes were used, with two routes used twice for a total of 6 trials per condition (single and dual-task, with and without previewing), and 24 trials in total. All participants walked the same routes in a pre-set order but never walked the same-coloured route on consecutive trials. Participants were asked to follow a designated coloured path at their preferred walking speed, contacting each target with any part of their foot, but without needing to step precisely on its centre (as in: Cocks et al, 2025). A video camera positioned at the end of the walkway recorded all trials to allow offline coding of stepping errors and cross-steps.

**Figure 1.**
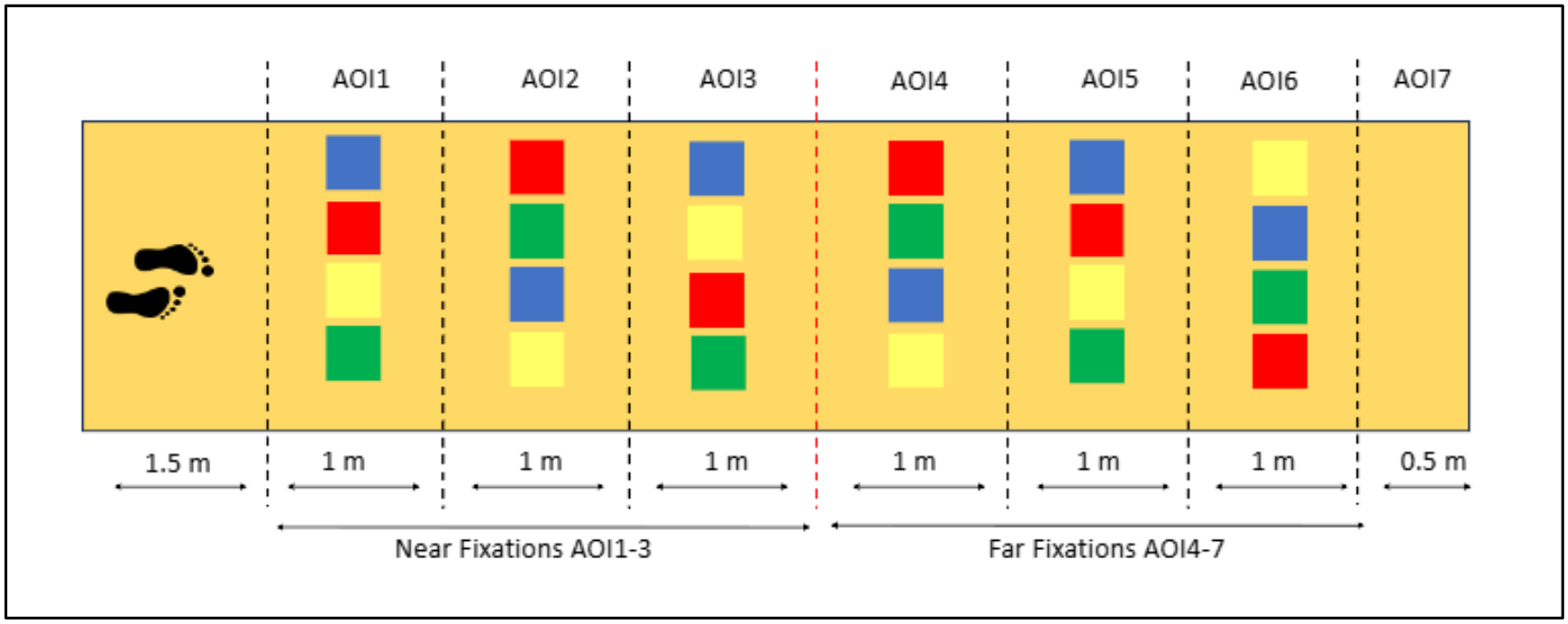
Schematic representation of the adaptive walking route. The walkway was divided into seven task-relevant Areas of Interest (AOIs 1-7), extending from proximal to distal (AOI 4-7) locations along the route. The red dashed line indicates the boundary between near and far AOIs. AOI 8 (white) any task-irrelevant areas outside the walkway.

Participants wore mobile eye-tracking glasses (SensoMotoric Instruments, Teltow, Germany) that sampled gaze position at 50 Hz. A scene camera integrated into the frame recorded the participant’s field of view at 25 Hz with a resolution of 1920 × 1080 pixels. Prior to each experimental condition, a three-point calibration was performed. Calibration quality was subsequently verified between trials using a visual target check, with recalibration undertaken if accuracy was deemed insufficient. The walkway was divided into seven task-relevant Areas of Interest (AOIs) spanning the near to distal regions of the route, with an eighth AOI representing task-irrelevant areas outside the walkway (see Fig. 1). Task relevant AOIs included in the analysis contained the stepping targets and were used to quantify visual sampling behaviour during route previewing. For analysis purposes, AOIs 1-3 were classified as near fixations, representing immediately upcoming stepping constraints and the 2/3 step look-ahead window previously described by Patla and Vickers (2003) and Matthis & Fajen (2014). AOIs 4-7 were therefore classified as far fixations, representing future route information (see Fig. 1). Fixations were identified using the SMI automated dispersion-based algorithm which identified fixations as groups of consecutive points within a maximum separation of 1 degree of the visual angle, and automated minimum fixation duration of 80ms (Salvucci & Goldberg, 2000) and each fixation was assigned to an AOI for subsequent analysis.

### 2.3 Walking and Dual-Task Procedure

Participants completed adaptive walking trials under single-task and cognitive dual-task conditions in a set order (McVeigh-Mellor et al., 2026). In dual-task trials, participants performed verbal serial subtraction aloud in steps of seven, beginning from a random three-digit number (100-300) (Hausdorff et al., 2008). A seated 30-second subtraction baseline was completed separately at the end of the walking conditions to compute cognitive dual-task cost. The correct number of responses was divided by trial duration, to determine the correct response rate per second. This allowed direct comparison of cognitive task performance during walking trials and the baseline.

In previewing conditions, participants were first instructed which coloured route to follow while facing away from the walkway. Once told “begin previewing now” participants turned 180° to face the walkway and initiated a 10 second stationary previewing phase (timed using a stopwatch, starting once they had completed their turn and were facing the walkway; based on Curzon-Jones and Hollands (2018). The participants were not explicitly instructed to preview the route in any specific way but simply told to use the 10 seconds to look at the route they would be walking, allowing individuals to spontaneously adopt their own previewing strategy. Following a “Stop previewing” instruction, they turned away from the walkway, received the serial subtraction number to start from (dual-task only) followed by a “Go” cue to commence walking and counting. This ensured that previewing occurred before any cognitive load was introduced. The same procedure (bar the use of the dual-task instruction) was used for single-task conditions to ensure the turning away and back to the walkway procedure was consistent across conditions.

### 2.4 Outcome measures

#### 2.4.1 Data Preparation

We only included data from (i) participants who had sufficient data available for ≥4 trials in the dual-task previewing condition and only included (ii) trials for which a minimum of 50% of the identified fixations had position data (Ellmers et al., 2016). In total, 10 participants from the original dataset (N=39) were therefore excluded from the analysis; three participants were excluded due to complete failure to capture eye-tracker data due to operator error or calibration failure. For the remaining 7 participants, fixation position could not be identified for >50% of trial duration of >4 trials, leaving 29 participants’ data to be analysed.

#### 2.4.2 Gaze Variables

Only task-relevant fixations (fixations to the walkway) were included in the analysis of the gaze behaviour during the previewing phase. We used Lempel-Ziv complexity (LZC_task) to quantify the complexity of each participants’ gaze behaviour by measuring how many patterns were encountered within the given previewing period (Lempel & Ziv, 1976). To compute LZC_task, AOI fixation sequence was parsed sequentially, each time a new subsequence appeared it was added to an index of possible patterns, with the total number of unique patterns reflecting the sequence’s complexity (Lounis et al., 2020). This count was then normalised by sequence length to allow comparison across signals to summarise how varied the sequence of gaze locations was during the preview period. Higher LZC_task values indicate greater randomness or unpredictability, whereas lower values reflect a more repetitive structure (Harris et al., 2023). This measure has previously been used to distinguish differences in visual scanning behaviour between novice and experienced pilots (Lounis et al., 2020). The maximum AOI transition size was used to describe the largest shift in gaze between route regions. Larger transitions may indicate that participants display less granular visual sampling of the route. Maximum AOI transition size per trial (MaxAOI_Transition) was calculated as the largest change between consecutive fixations within the previewing phase (calculated as the change from one AOI to the next [e.g., a transition from AOI 1 to AOI4 would be noted as a change size of 3]). Distal allocation of gaze (FarFix) was quantified as the percentage of total fixation duration directed at the furthest areas (AOI 4-7) of the walkway (see Fig. 1). FarFix was selected as the primary fixation metric because it was considered to most directly reflect distal visual previewing. Greater percentage of fixation duration beyond the immediate walkway was expected to reflect the acquisition of information relevant to future stepping actions.

#### 2.4.3 Performance variables

Walking and cognitive performance were assessed using four dependent variables. Walking speed (m/s per trial), step errors (number of missed route targets per trial), cross-step proportion (rate of cross-steps per trial, calculated as number of cross-steps divided by number of total steps). Cross-steps were defined as steps in which one foot crossed the midline or lateral border of the contralateral stance foot (Cocks et al., 2025) and are considered a potential marker of unsafe stepping strategies particularly for older adults at risk of falls (Cocks et al., 2025). Cross-step behaviour was analysed using a two-part approach: firstly, a binary measure (cross-step occurrence) indicating whether a cross-step occurred in a given trial, and (ii) a continuous cross-step proportion measure for trials in which cross-stepping occurred. Motor (speed) and cognitive dual-task cost (DTC) were included as a measure of performance during walking, cognitive DTC was measured as proportional change in response rate from seated single-task baseline serial subtraction to dual-task waking with serial subtraction in the previewing condition (Single-task - Dual-task preview) / Single-task x 100 per trial) (Montero-Odasso et al., 2012).

#### 2.4.4 Statistical Analysis

All analyses were conducted in MATLAB (MathWorks, Natick, MA) using restricted maximum likelihood estimation for linear mixed-effects models and maximum pseudo-likelihood estimation for generalized linear mixed-effects models. Statistical significance was set at *p* < .05.

Linear mixed-effects modelling was used to examine associations between previewing gaze behaviour and adaptive walking performance outcomes in the dual-task previewing condition. Mixed-effects approaches were selected because the dataset contained repeated trial-level observations nested within participants, requiring models capable of accounting for within-subject dependency while retaining trial-level variability. Participant was therefore included as a random intercept in all models.^1^

Cross-step behaviour was analysed using a two-part modelling approach. Initially, to understand if these gaze-related variables were associated with the *emergence* of this stepping strategy, cross-step occurrence was examined using a generalized linear mixed-effects model with a binomial distribution and logit link function, where trials were coded according to whether a cross-step occurred (0 = no cross-step, 1 = cross-step present). Subsequently to further probe the strength of this association the occurrence frequency of cross-steps was examined using a linear mixed-effects model restricted to trials in which cross-stepping occurred, to investigate whether previewing variables predicted the frequency of cross-step behaviour.

Separate linear mixed-effects models were constructed for walking speed, step errors, and cognitive DTC. Cognitive DTC values (n=6) exceeding ±300% were excluded prior to analysis because these values were considered likely to reflect very small single-task denominators rather than meaningful behavioural variation. A sensitivity analysis retaining these extreme values was conducted to assess whether their exclusion materially affected the conclusions.

## 3. Results

### 3.1 Participants

Characteristics of the 29 participants that were included in this analysis are presented in Table 1. Participants were healthy older adults with good mobility and cognitive function, with a mean Timed Up-and-Go time of 8.0 s (SD = 1.4) and MoCA score of 26.9 (SD = 2.2).

**Table 1.** Participant Demographics.

| **Demographics** | **Mean (SD)** | **Range** |
|---|---|---|
| **Age** | 73.9 (8.1) | 60-87 |
| **Sex (Male/Female)** | 10/19 | n/a |
| **Height (cm)** | 162.0 (13.0) | 142-182 |
| **Weight (kg)** | 74.7 (16.6) | 43-110 |
| **Visual Acuity\*** | 20/25 | 20/20–20/40 |
| **Timed-up-and-Go (seconds)** | 8.0 (1.4) | 5-11 |
| **Montreal Cognitive Assessment (0-30)** | 26.9 (2.2) | 23-30 |
| **Falls Efficacy Scale - International (16-64)** | 22.8 (6.0) | 16-42 |
| **Participants with ≥1 fall in previous year (n)** | 8 | n/a |
| **Updated Perceived Control over Falling (0-20)** | 16.9 (2.8) | 12-20 |
| **Education (High school/University/post-grad/other) (N)** | 8/12/5/4 | n/a |

*Note \* Presented as median*

### 3.2 Previewing gaze characteristics

Descriptive statistics for previewing gaze characteristics and associated gait outcomes from the dual-task previewing condition are presented in Table 2.

**Table 2.**
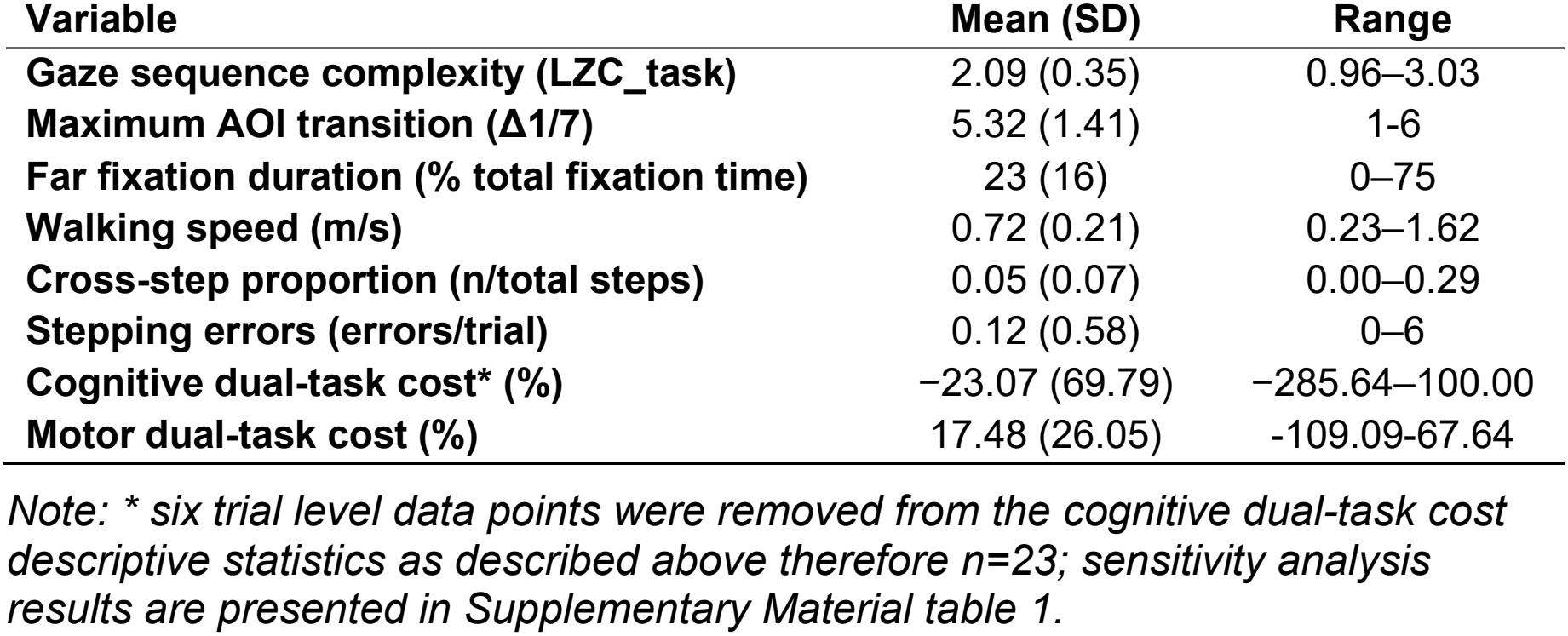
Previewing phase gaze characteristics and associated gait outcomes.

| **Variable** | **Mean (SD)** | **Range** |
|---|---|---|
| **Gaze sequence complexity (LZC_task)** | 2.09 (0.35) | 0.96–3.03 |
| **Maximum AOI transition (Δ1/7)** | 5.32 (1.41) | 1-6 |
| **Far fixation duration (% total fixation time)** | 23 (16) | 0–75 |
| **Walking speed (m/s)** | 0.72 (0.21) | 0.23–1.62 |
| **Cross-step proportion (n/total steps)** | 0.05 (0.07) | 0.00–0.29 |
| **Stepping errors (errors/trial)** | 0.12 (0.58) | 0–6 |
| **Cognitive dual-task cost\* (%)** | −23.07 (69.79) | −285.64–100.00 |
| **Motor dual-task cost (%)** | 17.48 (26.05) | -109.09-67.64 |

*Note: \* six trial level data points were removed from the cognitive dual-task cost descriptive statistics as described above therefore n=23; sensitivity analysis results are presented in Supplementary Material table 1.*

Table 3 presents the linear mixed-effects models examining whether previewing gaze behaviour predicted gait and cognitive outcomes. Larger maximum AOI transitions were associated with slower walking speed (β = −1.01, SE = 0.19, *p* < .001), and greater dual-task-related reductions in walking speed (as evidenced by positive associations with speed dual-task cost, β = 1.01, SE = 0.19, *t* = 5.32, *p* < .001). Greater gaze sequence complexity (LZC_task) was associated with lower cognitive dual-task cost (β = −34.39, SE = 16.18, *p* = .035; Figure 2a). No significant associations were observed between any of the previewing gaze variables and step errors (all *p* > .464).

**Table 3.**
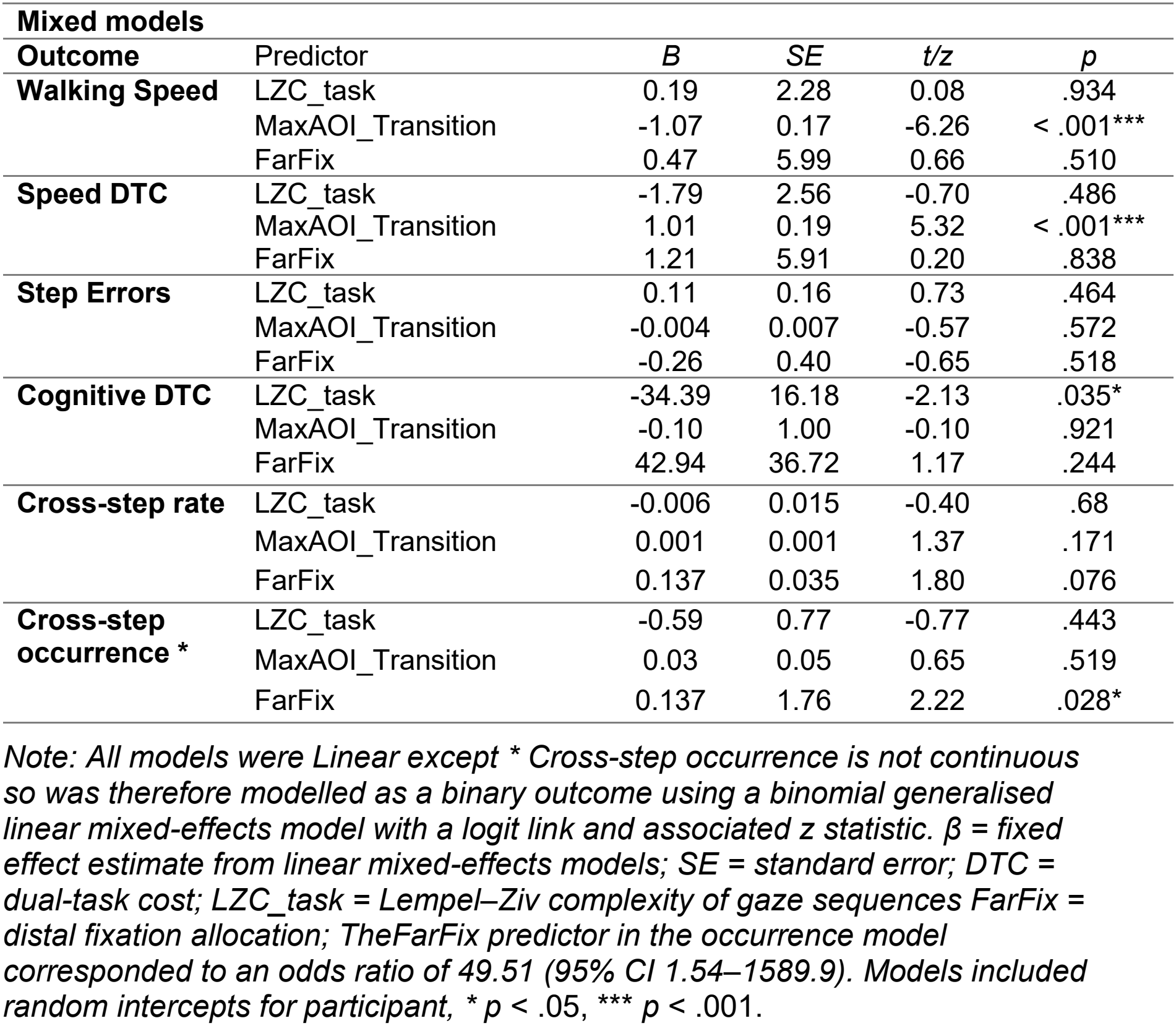
Mixed-effects models examining associations between previewing variables and gait and cognitive outcomes during adaptive walking.

| **Mixed models** |  |  |  |  |  |
| **Outcome** | Predictor | *B* | *SE* | *t/z* | *p* |
|---|---|---|---|---|---|
| **Walking Speed** | LZC_task | 0.19 | 2.28 | 0.08 | .934 |
|  | MaxAOI_Transition | -1.07 | 0.17 | -6.26 | < .001*** |
|  | FarFix | 0.47 | 5.99 | 0.66 | .510 |
| **Speed DTC** | LZC_task | -1.79 | 2.56 | -0.70 | .486 |
|  | MaxAOI_Transition | 1.01 | 0.19 | 5.32 | < .001*** |
|  | FarFix | 1.21 | 5.91 | 0.20 | .838 |
| **Step Errors** | LZC_task | 0.11 | 0.16 | 0.73 | .464 |
|  | MaxAOI_Transition | -0.004 | 0.007 | -0.57 | .572 |
|  | FarFix | -0.26 | 0.40 | -0.65 | .518 |
| **Cognitive DTC** | LZC_task | -34.39 | 16.18 | -2.13 | .035* |
|  | MaxAOI_Transition | -0.10 | 1.00 | -0.10 | .921 |
|  | FarFix | 42.94 | 36.72 | 1.17 | .244 |
| **Cross-step rate** | LZC_task | -0.006 | 0.015 | -0.40 | .68 |
|  | MaxAOI_Transition | 0.001 | 0.001 | 1.37 | .171 |
|  | FarFix | 0.137 | 0.035 | 1.80 | .076 |
| **Cross-step occurrence \*** | LZC_task | -0.59 | 0.77 | -0.77 | .443 |
|  | MaxAOI_Transition | 0.03 | 0.05 | 0.65 | .519 |
|  | FarFix | 0.137 | 1.76 | 2.22 | .028* |

*Note: All models were Linear except \* Cross-step occurrence is not continuous so was therefore modelled as a binary outcome using a binomial generalised linear mixed-effects model with a logit link and associated z statistic. β = fixed effect estimate from linear mixed-effects models; SE = standard error; DTC = dual-task cost; LZC\_task = Lempel–Ziv complexity of gaze sequences FarFix = distal fixation allocation; TheFarFix predictor in the occurrence model corresponded to an odds ratio of 49.51 (95% CI 1.54–1589.9). Models included random intercepts for participant, \* p* < .05, *\*\*\* p* < .001.

**Figure 2.**
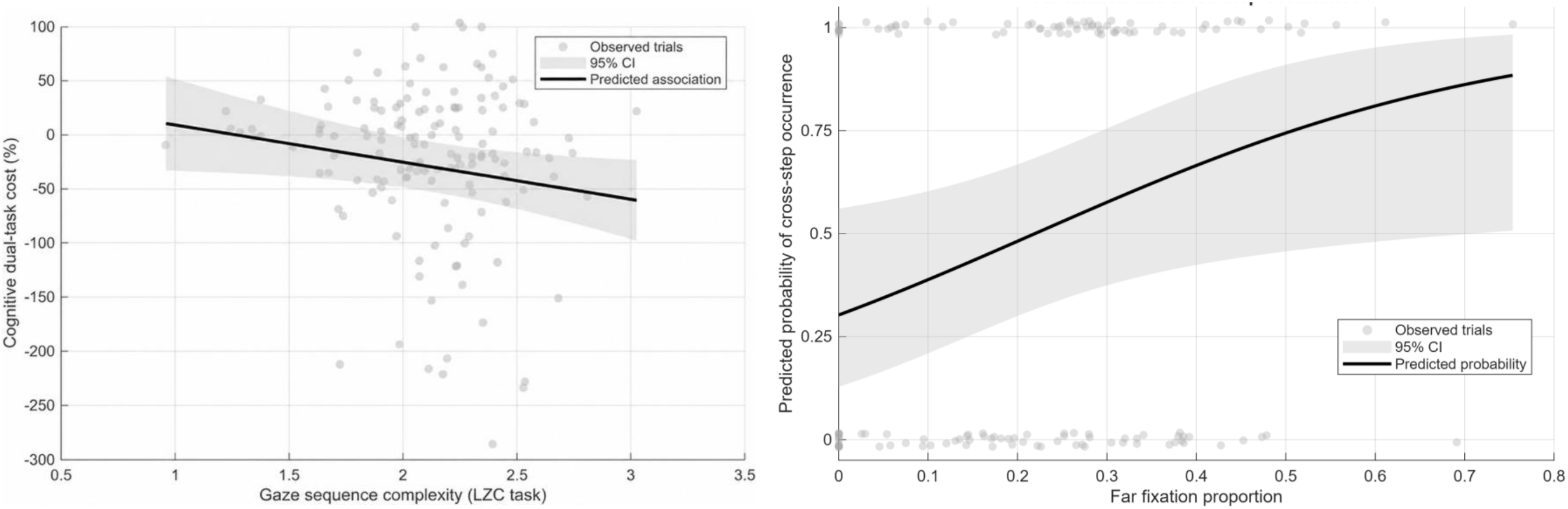
Association between gaze sequence complexity during previewing (LZC_task) and cognitive dual-task cost during adaptive walking. Higher LZC_task values reflect greater gaze sequence complexity and negative dual-task cost indicate improved performance.

**Figure 3.** Predicted probability of cross-step occurrence as a function of distal fixation allocation (FarFix) during the previewing phase, FarFix reflects the proportion of fixation duration directed toward distal walkway regions during previewing. *Note: The solid line represents model*-*estimated probabilities derived from a generalized linear mixed*-*effects model. The shaded band indicates the 95% confidence interval. Grey points represent observed trial*-*level data. Models included random intercepts for participant*.

### 3.3 Gaze behaviour and stepping strategy

Greater distal fixation allocation (FarFix) was significantly associated with increased likelihood of cross-step occurrence, β = 0.137, SE = 1.76, p = .028 (see Fig. 2b). Among trials in which cross-stepping occurred, FarFix also demonstrated a positive, but non-signficant, association with cross-step rate, β = 0.137, SE = 0.035, p = .076. No significant effects were observed for LZC_task or MaxAOI_Transition in either model (all p > .443; see Table 4 for full results

## 4. Discussion

Previous work has shown that route previewing can reduce dual-task related gait impairments in older adults (McVeigh-Mellor et al., 2026). The present analysis explored the gaze characteristics underlying these behavioural benefits by examining which specific gaze behaviours during previewing predict subsequent gait and cognitive performance under dual-task conditions. We hypothesised efficient visual exploration, characterised by smaller maximal gaze transitions and greater relative proportion of distal fixation duration towards the latter half of the walkway, would support feedforward planning and improve subsequent walking performance. Further, gaze-sequence complexity was explored as a novel indicator of visual planning whereby greater complexity was predicted to be associated with worse walking performance.

Notably, larger maximal gaze shifts (i.e. greater transition size of consecutive fixations between AOIs) predicted slower walking speed, but were unrelated to stepping errors or cross-stepping. This suggests that making larger gaze shifts during previewing may reduce the efficiency of route encoding before movement begins. Larger transition size also predicted greater walking speed dual-task costs, indicating that larger gaze shifts during previewing were associated with greater dual-task-related reductions in subsequent walking speed. Participants who shifted gaze between widely separated areas of interest may have encoded the route as a series of isolated stepping locations rather than as a continuous sequence of future foot placements, leading to a more cautious walking strategy (Boschker et al., 2002). This interpretation is consistent with evidence that the organisation of visual search can be behaviourally meaningful. Amor et al. (2016) examined the temporal organisation of eye movements and found that participants adopted a systematic reading-like scanning strategy. Therefore, *how* gaze progresses between locations, may provide information about the organisation of visual exploration, beyond simply where fixations are directed. More specifically within locomotion research, effective feedforward control has been shown to depend on sampling terrain within an optimal look-ahead window of approximately two step lengths (Patla & Vickers, 2003; Matthis, 2018). If route previewing facilitates the formation of action-relevant chunks (Boschker et al., 2002), large transitions between distant route locations may disrupt the sequential organisation of these chunks, leading to slower subsequent walking speed as previously reported (McVeigh-Mellor et al., 2026). However, maximum transition size alone does not provide information about the broader organisation of visual exploration. In this context a large transition may also reflect a purposeful shift to inspect distal route features before returning to more proximal locations. Consequently, caution is warranted when interpreting larger transitions as inherently less efficient. It is possible that the benefits of previewing depend less on the availability of preview and more specifically on how route information is encoded into a prospective movement plan before gait initiation (Pezzulo et al., 2010).

Among all gaze measures examined, only gaze-sequence complexity was associated with cognitive dual-task cost. This suggests that the organisation of visual exploration during previewing may be particularly relevant to the cognitive demands experienced once walking begins. In the present study, greater gaze sequence complexity (LZC_task) was associated with lower cognitive dual-task costs alone, not with differences in gait performance itself. These results may suggest that scan-path organisation primarily influenced the cognitive resources required for route retention rather than the direct control of stepping movements itself (Ellmers et al., 2016). Because working memory resources are shared across locomotor and cognitive processes, this benefit may have emerged primarily as reduced cognitive dual-task cost. A potential interpretation is that more complex visual exploration facilitated the construction of an internal route representation, potentially reducing the visuospatial working-memory demands associated with route retention and online planning during walking (Baddeley, 2003; Pezzulo et al., 2010). It is possible that sampling the route in a more complex manner supported gait planning/performance by reducing the online demands associated with this, thus increasing the relative attentional capacity available for the secondary cognitive task.

Contrary to our hypothesis, greater distal fixation allocation was not associated with improvements in gait or cognitive dual-task performance. Instead, greater distal previewing increased the likelihood of cross-step occurrence..This finding was unexpected; in our previous work we showed that the presence of a cognitive dual-task increased the frequency of cross-stepping, (McVeigh-Mellor et al., 2026). Therfore, we expected distal previewing to reduce the occurrence of cross-steps as an automatic stepping response when attentional resources were divided. This association between distan fixation and cross-step frequency suggests that the availability of advance visual information has a less distinct relationship with subsequent stepping behaviour.The current finding could be consistent with Cocks et al., (2025), who demonstrated that reduced cognitive resources increase reliance on risky automatic stepping strategies in older adults. However, participants who allocated a greater proportion of gaze towards distal route features during previewing remained more likely to adopt cross-steps, indicating substantial individual variability in how previewed information was incorporated into locomotor planning. Instead, in the context of previewing, looking further ahead may reflect the detection of perceived route challenge rather than more effective feedforward planning. This is not dissimilar with findings from climbing research demonstrating that visual exploration increases when participants encountered environmental features that approach the limits of their action capabilities, reflecting a search for functional movement solutions rather than straightforward route planning (Van Knobelsdorff et al., 2020). Pezzulo et al. (2010) proposed that successful previewing involves evaluating how each upcoming movement constrains the next, while Russo et al. (2025) demonstrated that performing serial-7 subtraction while walking reduced fixations on task-relevant stepping targets and restricted visual exploration between proximal and distal areas of the walkway suggesting that cognitive load may constrain the visual search processes required for feedforward gait planning. Together, these findings suggest that the behavioural benefits of previewing depend on efficiently integrating visual information across the upcoming route to support feedforward locomotor planning, rather than simply increasing attention towards more distant stepping locations.

An important observation from the present study was that no single gaze characteristic consistently predicted all behavioural outcomes. Instead, larger gaze transitions predicted walking speed, greater distal fixation allocation predicted cross-step occurrence, and gaze-sequence complexity predicted cognitive dual-task cost indicating that different aspects of previewing were selectively associated with different dimensions of adaptive walking. This pattern aligns with theoretical accounts of visually guided locomotion, which propose that vision supports multiple complementary functions including environmental sampling, route planning and movement regulation rather than a single visual strategy (Patla, 1997; Reynolds & Day, 2005; Matthis et al., 2017).

### 4.1 Limitations

Ten participants (25.6% of the recruited sample) were excluded from the present analysis due to insufficient eye-tracking data. Although data-quality-related participant loss is a recognised challenge in eye-tracking research (Holmqvist et al., 2023), the resulting reduction in sample size and potential for exclusion bias should be acknowledged. The Cognitive DTC findings were robust to the exclusion of extreme DTC values, as retaining these observations did not alter the overall pattern of statistical inference (supplementary data Table 1). However, coefficient estimates and standard errors were substantially inflated when extreme values were retained, supporting caution when interpreting percentage-based DTC measures where small single-task denominators can generate extreme values. Interpretation of fixation duration remains complex, as prolonged viewing may reflect either effective information acquisition or uncertainty-driven monitoring. The absence of associations between distal fixation duration (FarFix) and gait outcomes suggests that duration alone is not a reliable indicator of previewing efficiency in this context. Nieuwenhuys (2008) discussed how in the investigation of professional climbers more fixations did not necessarily mean better visual behaviour, instead it reflected reduced processing efficiency. This is consistent with evidence from visual expertise research indicating that successful performance depends less on the amount of time spent viewing particular locations than on selectively directing gaze towards task-relevant information (Gegenfurtner et al., 2011). Together, these findings suggest that the organisation and relevance of visual sampling may be more informative than fixation duration alone. Although, we cannot determine what information was acquired during preview or how participants used it while walking. The fixed route may also limit generalisability of findings, as it is unclear if these findings could apply to navigation tasks where participants can choose which route to follow (rather than this being prescribed).

Finally, this analysis focused solely on pre-walking previewing, it did not attempt to determine how previewed information interacts with online gaze behaviour during adaptive gait. Although the observational design limits causal inference, previous research has demonstrated that gaze training can modify visual search behaviour and improve aspects of motor performance in other populations (Wood et al., 2017; Gunn et al., 2019). These findings suggest that targeted gaze training may be feasible, although its effectiveness for improving adaptive gait in healthy older adults remains uncertain. Future research could investigate whether prompting older adults to systematically integrate proximal and distal route information during previewing improves subsequent stepping performance, particularly during cognitive loading conditions.

## 5. Conclusion

Overall, the findings show that different aspects of older adults’ visual previewing behaviour support different locomotor and cognitive outcomes. Importantly, higher gaze-sequence complexity was linked to lower cognitive dual-task cost, suggesting that flexible and information-rich visual exploration may increase availability of cognitive resources during subsequent locomotion. Larger gaze transition sizes during previewing predicted slower walking speed, while greater duration of distal fixation allocation was associated with an increased likelihood of cross-step occurrence. Together, these findings may indicate that effective previewing depends not simply on sampling more distant route information (i.e. planning further ahead), but on integrating proximal and distal route information into a useful sequence of upcoming stepping actions. Large shifts between areas of interest, or preferential allocation of gaze towards distal areas, may be less beneficial when this information is not integrated with the intervening step/path information. When considered alongside previous findings that route previewing reduces dual-task costs, understanding how older adults visually plan upcoming walking routes may help inform interventions aimed at improving adaptive walking and reducing dual-task-related adaptive walking performance.

## Data Availability

All data produced in the present study are available upon reasonable request to the authors

https://doi.org/10.5281/zenodo.24286232

## 6. Acknowledgements

The authors would like to thank MSc students Emma Vincent, Yi Sum Agatha Siu, Sy Yue, Giuditta Simio, Yasser Bader, and Owen Lin for their assistance with data collection and the Brunel Older Adults Reference Group members for their participation.

## 7. Supplementary data

Supplementary data associated with this article can be found at https://doi.org/10.5281/zenodo.24286232

## 9. Declaration of use of generative AI and AI-assisted technologies

No generative AI or AI-assisted technologies were used in the creation, writing, revision, or preparation of this manuscript. All work was conducted entirely by the authors.

## 10. Author contributions: CRediT

**Olivia McVeigh Mellor:** Conceptualization, Methodology, Investigation, Data Curation, Formal Analysis, Visualization, Writing – Original Draft, Writing – Review & Editing **Adam J. Cocks:** Conceptualization, Methodology, Investigation, Writing – Review & Editing, Supervision **Liam O’Reilly:** Writing – Review & Editing **Dominic Orth:** Writing – Review & Editing **Adam Bruton:** Writing – Review & Editing **Toby Ellmers:** Writing – Review & Editing **Elmar Kal:** Conceptualization, Methodology, Investigation, Writing – Review & Editing, Supervision

## Footnotes

1 We originally planned multiple linear regression analyses but subsequently opted for linear and generalised linear mixed-effects modelling to account for repeated trial-level observations nested within participants and retain trial-level variability. The previewing gaze variables and cross-step analysis were also refined to better represent the visual sampling and behavioural outcomes of interest. This document (10.5281/zenodo.24286764) outlines all changes to the methods and analyses previously registered.

